# Childhood malnutrition and bacteraemia at a tertiary hospital in Malawi

**DOI:** 10.1101/2021.08.19.21262269

**Authors:** Victoria Temwanani Mukhula, Philliness Prisca Harawa, Chisomo Phiri, Stanley Khoswe, Jessica Chaziya, Emmie Mbale, Caroline Tigoi, Pui-Ying Iroh Tam, Robert Bandsma, Wieger Voskuijl

## Abstract

**Background:** Malnutrition increases risk of acquiring infections but clinical characteristics and hospital outcomes among children in low resource settings with high rates of antimicrobial resistance have not been clearly described.

**Aim:** Our main aim was to ascertain prevalence of bacteraemia in hospitalised children at Queen Elizabeth Central Hospital, Malawi.

**Methods:** We conducted a secondary analysis of a prospective study of children who had a blood culture collected during hospitalisation.

**Results:** Out of 175 children who had blood cultures collected during hospitalisation, 75 had severe acute malnutrition (SAM), 31 moderate acute malnutrition (MAM), and 69 no acute malnutrition (NAM). Twelve (7%) had bacteraemia (8 SAM, 1 MAM, 3 NAM) and seventeen (10%) died (14 SAM, 2 MAM, 1 NAM). Fever, vomiting and convulsions were least common in SAM (69%, 37%, 1%) compared to MAM (90%, 81%, 10%) and NAM (99%, 46%, 29%; p<0.001) children. Mortality was significantly higher in those with than without bacteraemia (33% vs 8%, p=0.004). Most common isolates were *Salmonella Typhimurium* (31%) and *Escherichia coli* (23%). High rates of bacterial resistance were noted to gentamicin (58%), a first-line antibiotic, and ceftriaxone (33%), a second-line antibiotic.

**Conclusions:** Mortality and bacteraemia rates are highest in hospitalised children with SAM. Despite this, SAM children do not present with typical clinical features, including fever, vomiting and convulsions. Given the high rate of antimicrobial resistance in this setting, a high index of infection clinical suspicion, awareness of local susceptibility patterns and evidence-based antibiotic guidelines are needed to optimise clinical care and antimicrobial stewardship.

**Lay Summary:** Malnutrition increases the risk of having an infection but symptoms and hospital outcomes among children with malnutrition, in countries like Malawi with high rates of antimicrobial resistance, have not been clearly described. This study describes a study of children who had a blood culture collected during admission to Queen Elizabeth Central Hospital, Malawi. Of 175 children who had blood cultures collected, 12 (7%) had a bacteria found (‘bactaeriemia’) and 17 (10%) died. Fever, vomiting and convulsions were significantly less common in severe malnutrition compared to children with moderate malnutrition and those with no malnutrition. Mortality was significantly higher in those with bacteraemia than without. High rates of bacterial resistance were noted to first- and second-line antibiotics. Mortality and bacteraemia rates are highest in hospitalised children with SAM even though they do not present with typical features of bacteraemia.

## Introduction

Malnutrition is estimated to contribute up to 45% of all childhood mortality globally, the majority of which are in low- and- middle income countries (1,2). There is a bi-directional relationship between malnutrition and infection. Chronic or recurrent infections, like untreated HIV and diarrhoea, can often contribute to malnutrition. Conversely, due to impaired immune function, malnourished children are at increased risk of infectious diseases, including persistent diarrhoea, pneumonia and bacteraemia (3).

Bacteraemia is defined as the presence of viable bacteria in the circulating blood (4) and diagnosis is confirmed by blood culture. Prevalence of bacteraemia in malnourished children in Africa ranges between 5%-36% (5–7). Malnourished children with bacteraemia have an increased risk of poor outcomes including mortality and prolonged hospital stay (8).

Diagnosis of bacteraemia is challenging in low- and- middle income countries due to limited laboratory capacity. Apart from that, malnourished children do not usually display obvious clinical signs of infections like fever (9,10). The World Health Organization (WHO), therefore, recommends empirical treatment with broad spectrum antibiotics for all admitted severely malnourished children to prevent mortality (11). Concurrently, there has been a global increase in the prevalence of antimicrobial resistance in young hospitalised children, irrespective of their nutritional status, which has also been associated with poor clinical outcomes (13).

There is limited recent data evaluating the association between bacteraemia and outcomes in malnourished children, a high risk population (5,11–15). There are no data in Malawi on the prevalence of bacteraemia and its associated outcomes for hospitalised malnourished children. Therefore, we collected data on culture and susceptibility profiles in the hospitalised population to inform local Malawian guidelines on rational use of antibiotics among children with malnutrition.

The main objective of this study was to ascertain prevalence of bacteraemia in hospitalised children enrolled in the “Childhood Acute Illness and Nutrition network” (CHAIN-network) cohort study at Queen Elizabeth Central Hospital (QECH) in Malawi, a tertiary hospital which follows WHO recommendations on antimicrobial use in treating malnourished children. The secondary objectives were to determine the blood culture isolates and their susceptibility profiles; and compare clinical characteristics and outcomes among children with and without bacteraemia.

## Methodology

### Setting

The study took place at QECH which is the largest referral hospital located in Blantyre, southern Malawi. It serves an estimated population of 800,000 (16). The paediatric department admits 26,000 patients per year with approximately 1,200 patients served in the nutrition rehabilitation unit (17,18).

### Study design

The CHAIN Network is a collaborative group of investigators which was established in four African (Burkina Faso, Uganda, and Kenya) and two South Asian countries (Bangladesh, Pakistan) to improve child survival and optimise growth and development outcomes of malnourished children in low-resource settings. The main aim of CHAIN was to identify modifiable risk factors that could be targeted by interventions for children at highest risk of mortality, including malnourished children (19).

We conducted a secondary analysis of the CHAIN data from the Malawi site (19). In Malawi, CHAIN recruited hospitalised children aged between 7 days and 24 months between 1 October 2016 and 31 January 2019, and followed them up to 180 days after discharge. Patients were followed during the initial admission and subsequent re-hospitalisations to QECH. Children with a known congenital syndrome, cleft palate, known congenital cardiac disease, known terminal illness e.g. cancer, admitted mainly for surgery or trauma, or likely to require surgery within 6 months were excluded. The CHAIN cohort study collected extensive clinical and follow-up data of all participants.

Our study was a convenience sample where we included all children who were recruited in the CHAIN study and had a blood culture taken during admission, hospital stay or follow-up. We excluded children who did not have a blood culture collected. Written informed consent was provided, and approval was obtained from College of Medicine Research Ethics Committee (COMREC approval number: P.06/16/1969) under Kamuzu University of Health Sciences then called University of Malawi.

### Data collection

We collected data from the CHAIN’s Research Electronic Data Capture (REDCap) version 8. Clinical data included anthropometry, admission symptoms and co-morbidities, initial diagnosis, initial treatment received, and hospital stay. Children were stratified by mid upper arm circumference (MUAC) into three nutritional strata: 1) No Acute Malnutrition (NAM), defined as MUAC of >12.5 cm for children aged at least 6 months or >12 cm for those under 6 months of age; 2) Moderate Acute Malnutrition (MAM), defined as MUAC of 11.5-12.5 cm for children aged at least 6 months, or 11-12cm for <6 months of age; and 3) Severe Acute Malnutrition (SAM), defined as MUAC of <11.5 cm for those at least 6 months or <11cm for <6 months of age, or presence of nutritional oedema regardless of MUAC.

Corresponding blood cultures and antimicrobial susceptibility results were extracted from the Malawi-Liverpool Wellcome Trust laboratory system, following microbiological testing procedures described in detail elsewhere (20). Blood culture results were described as positive if there was significant growth of a pathogen, and negative if there was either a contaminant or no growth. The following isolates were considered to be contaminants: coagulase-negative *Staphylococci, Micrococcus* species, *Bacillus* species and diptheroids. Isolates from blood cultures taken after 48 hours of hospital stay were regarded as nosocomial infections while those from within 48 hours of admission were considered to be community-acquired infections.

### Statistical analyses

We exported data into Microsoft Excel 2007 and analyzed in R Studio Version 1.2.5001. Mean, standard deviation (SD) and ranges were used for continuous variables and with an ANOVA p-value. Counts and percentages were used to describe categorical variables and a Chi-square test was used to compare the differences in the groups. Statistical significance was set at P <0.05 with a confidence interval of 95%. Primary outcome was bacteraemia.

## Results

A total of 347 children were enrolled in the CHAIN study and 175 had blood cultures collected during admission: 75 with SAM, 31 MAM and 69 NAM (Figure 1).

**Figure 1.**
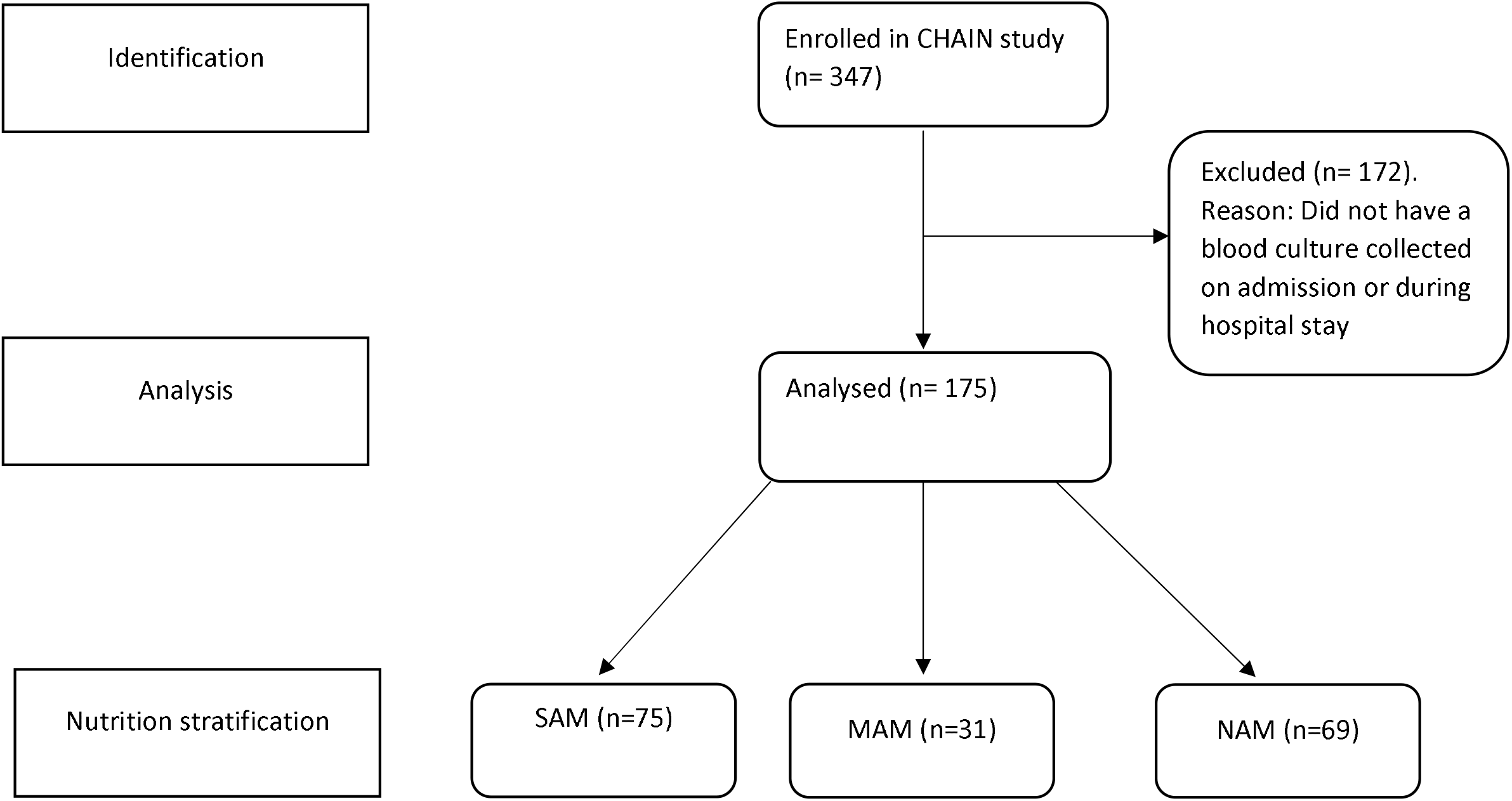
Participants’ flow diagram. MAM, Moderate Acute Malnutrition; NAM, No Acute Malnutrition; SAM, Severe Acute Malnutrition

Table 1 compares clinical characteristics and outcomes among patients of different nutrition groups. Several significant differences were noted. Children with MAM were the youngest with a mean age of 10 months and NAM were oldest with mean age of 13 months (p=0.039). All nutrition groups had negative mean Weight-for-Height Z (WHZ) score: NAM: -0.6 SD, MAM: -2.0 SD, SAM: -3.2 SD. HIV prevalence confirmed by DNA PCR testing was highest in SAM children (9%) followed by MAM (1%) and none in NAM. Having fever on admission was least common in SAM (69%) compared to MAM (90%) and NAM (99%). Vomiting and convulsions were least common features among SAM (37% and 1%) compared to MAM (81% and 10%) and NAM (46% and 29%). Sepsis was the least documented admission clinical diagnosis in children with SAM compared to MAM and NAM at 29%, 65% and 46%, respectively (p=0.003). Thirteen patients had a positive blood culture (7%) with significant growth, which was highest among children with SAM (12%), followed by those with MAM and children with NAM (1% and 2%, respectively). All of thirteen patients with positive blood culture grew bacterial species except one malnourished patient who grew candida. Twenty-seven patients (15%) grew contaminants. Fever was a more common presentation in patients with a negative blood culture (86%) compared to those with a positive blood culture (62%; p=0.017), irrespective of their nutritional status. Oral rehydration was highest among children with SAM (43%) compared with MAM and NAM (6% and 0%, respectively; p<0.001). Children with SAM had significantly longer hospital stays compared to MAM and NAM with mean days of 10, 5 and 3, respectively (p<0.001). Mortality was also highest in children with SAM at 19% compared to 6% in MAM and 1% in NAM (p=0.002).

**Table 1.**
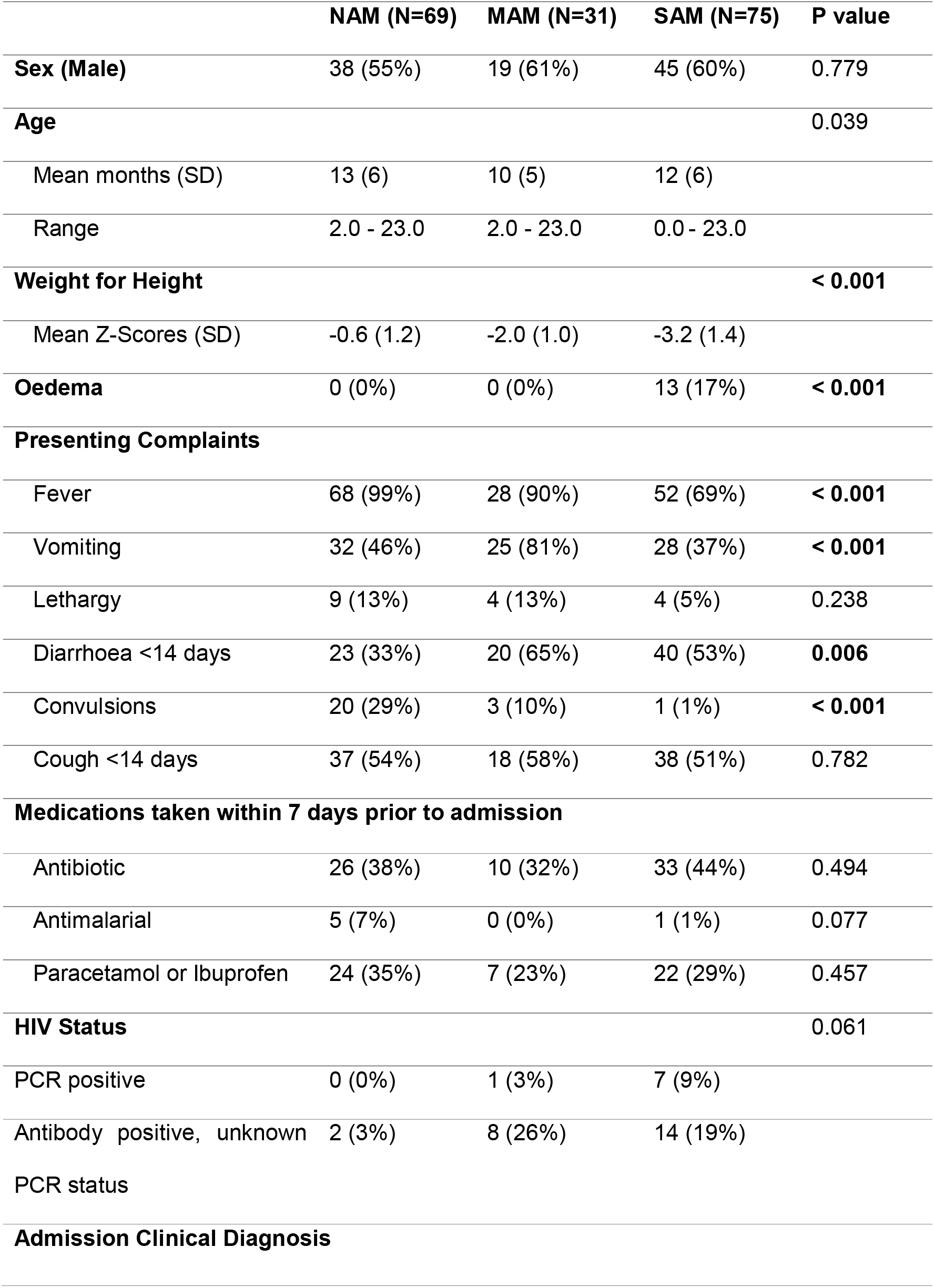
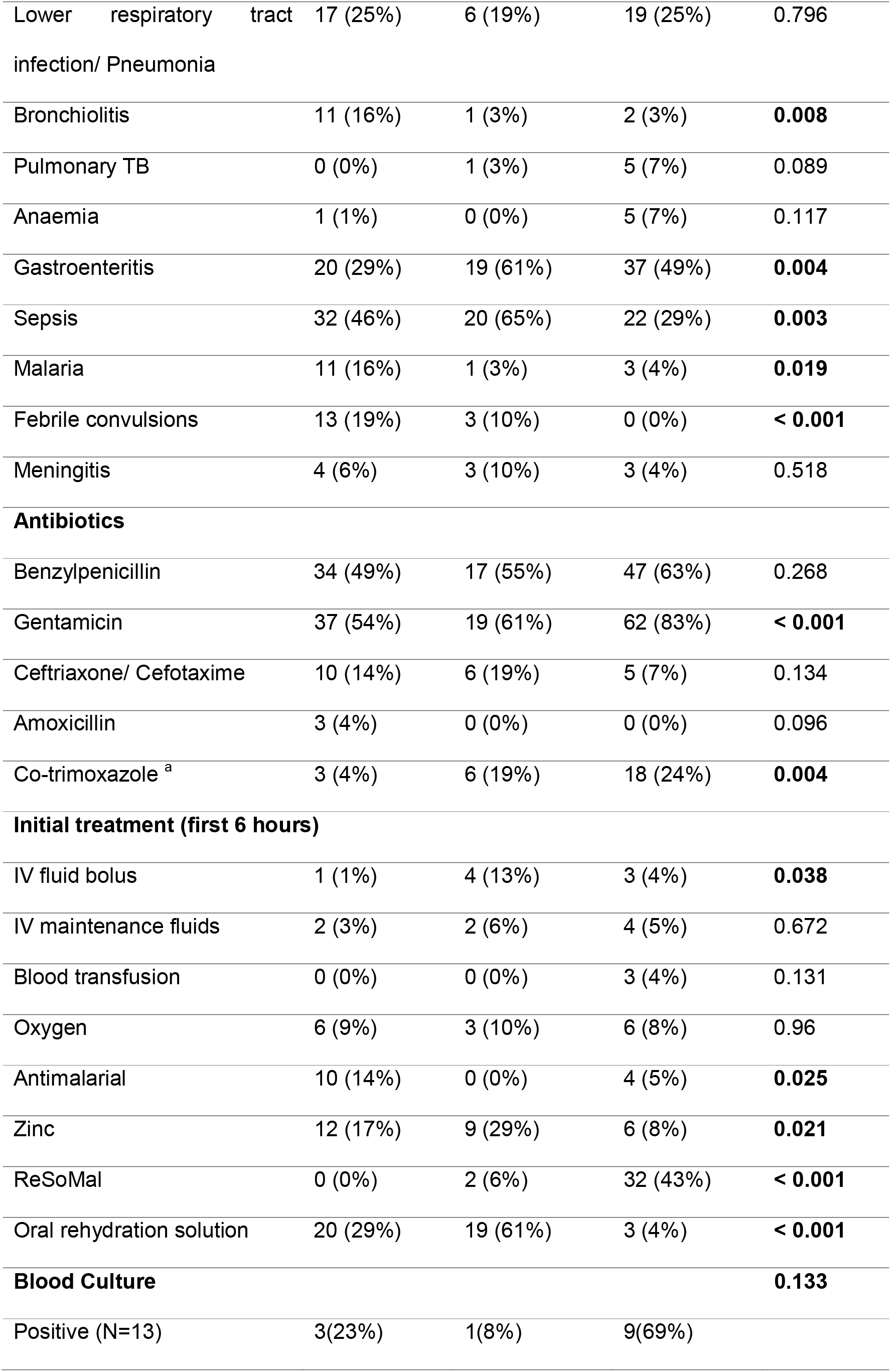
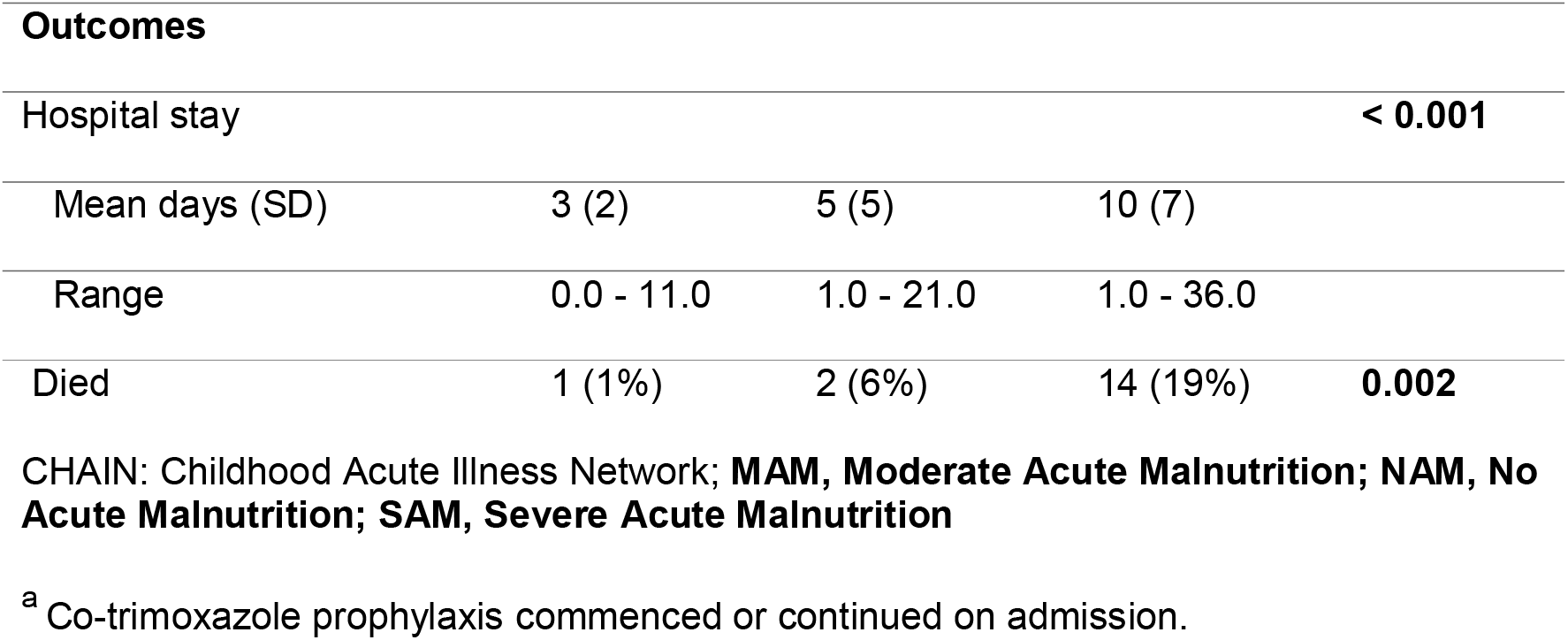
Comparison of clinical characteristics and outcomes among CHAIN patients who had blood culture obtained on admission, stratified by nutrition status

Table 2 describes characteristics and outcomes of patients with and without bacteraemia, shown through positive and negative blood cultures respectively. More patients with a positive blood culture received antibiotics within 7 days prior to admission than those with negative blood culture at 75% vs 37% (p=0.022). Patients with a positive blood culture had significantly higher mortality (33%) compared to those with a negative blood culture (8%, p=0.004). There were, however, no significant differences in nutrition status and duration of hospital stay between the two groups.

**Table 2.** Characteristics and outcomes of patients who had a positive and negative blood culture.

blood culture
| Blood culture | Negative <sup>a</sup> (N=163) | Positive (N=12) | P value |
| --- | --- | --- | --- |
| **Clinical characteristics** |  |  |  |
| Fever | 141 (87%) | 7(58%) | **0.009** |
| Diarrhoea (< 14 days) | 77 (47%) | 6(50%) | 0.853 |
| Poor feeding | 46 (28%) | 6 (50%) | 0.111 |
| Pneumonia | 39 (24%) | 3 (25%) | 0.933 |
| Sepsis | 69(42%) | 5(42%) | 0.964 |
| **Antibiotics in the past 7 days** | 60 (37%) | 9 (75%) | **0.009** |
| **Outcomes** |  |  |  |
| Hospital stay |  |  | 0.194 |
| Mean days (SD) | 6.1 (6.0) | 8.5(6.9) |  |
| Range | 0.0 - 36.0 | 2.0 - 26.0 |  |
| Died | 13 (8%) | 4 (33%) | **0.004** |

<sup>a</sup> No growth or non-significant growth or isolate other than bacteria

Of the 13 blood cultures that were positive,12 grew bacteria. *Salmonella Typhimurium* was the most common isolate (24%), followed by *E. coli* (18%). *Klebsiella pneumoniae* and *Candida* species were hospital acquired infections which were both isolated in children with SAM (Figure 2). Susceptibility profiles of these isolates to QECH paediatric first-line antibiotics (penicillin and gentamicin), second-line antibiotics (ceftriaxone and amikacin) and other commonly prescribed antibiotics (chloramphenicol and ciprofloxacin) are shown in Table 3. From the bacterial isolates, 58% were resistant to gentamicin, and 33% to ceftriaxone.

**Table 3.**
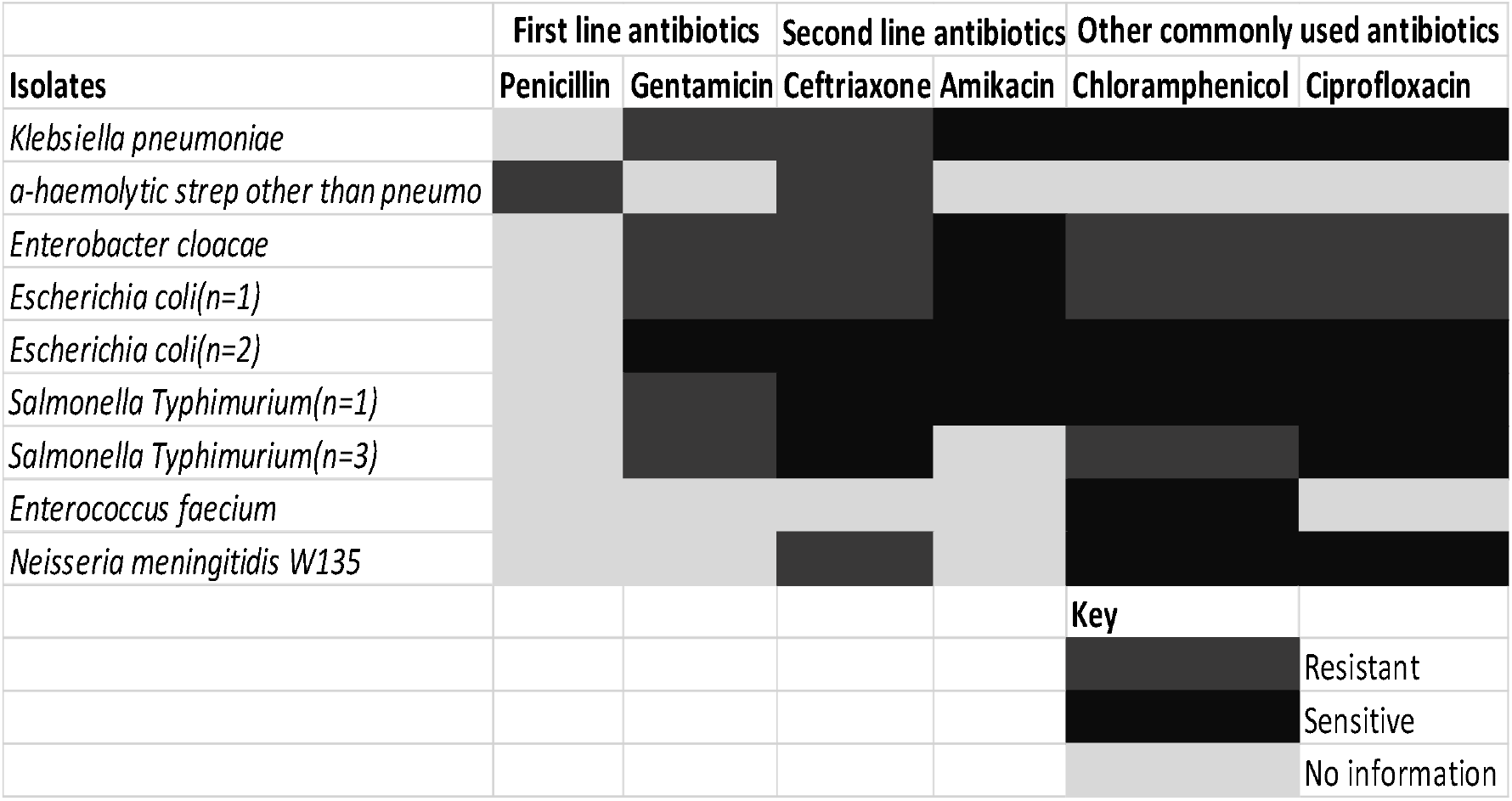
Sensitivity profiles of the blood culture isolates

**Figure 2.**
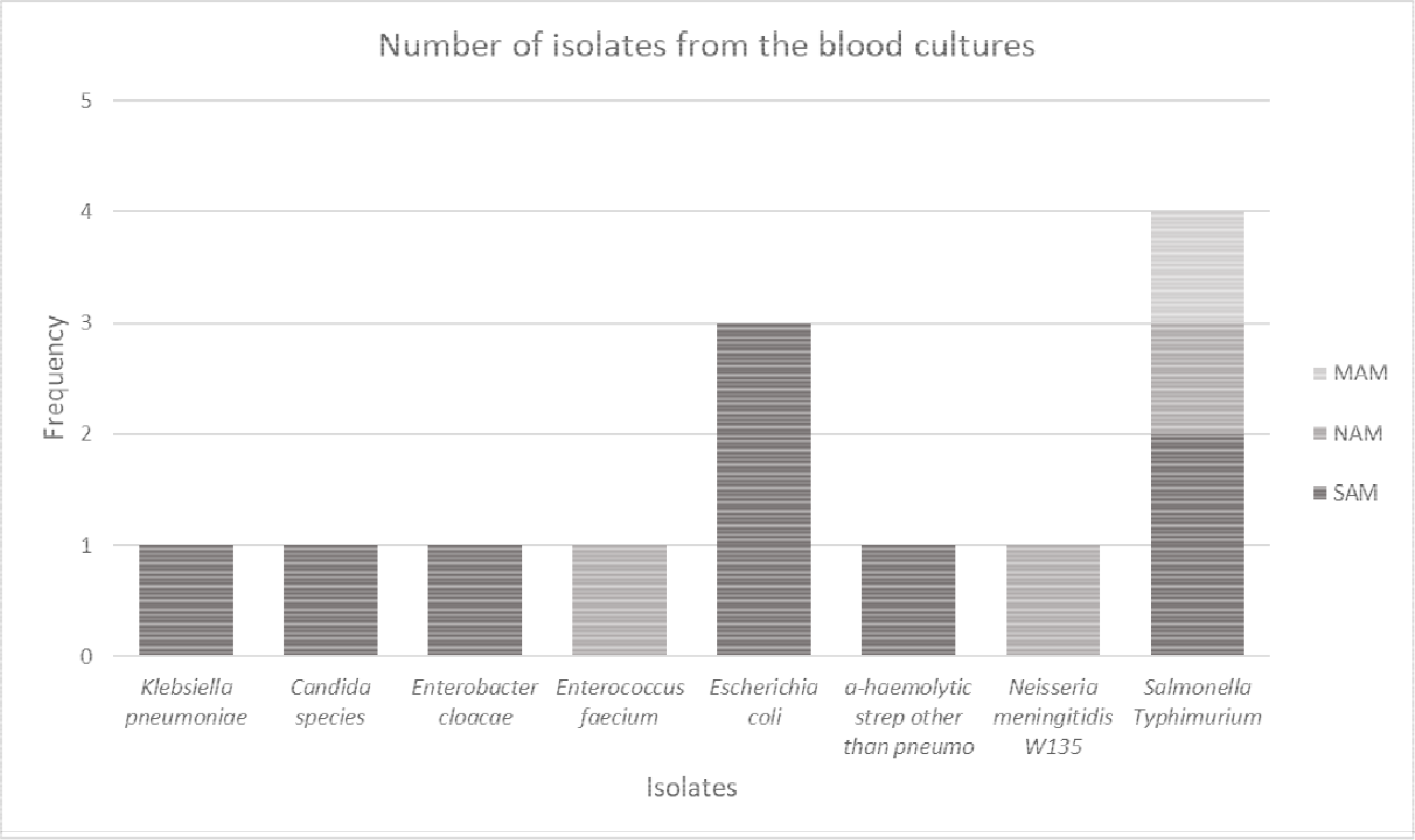
Isolates from different nutrition groups. MAM, Moderate Acute Malnutrition; NAM, No Acute Malnutrition; SAM, Severe Acute Malnutrition

Table 4 shows clinical characteristics of patients discharged compared to those who died. Overall, there was a 10% (n=17) mortality. Fever was a common clinical characteristic in those who were discharged (87%) compared to those who died (65%, p=0.017). Majority of the children who died presented with gastroenteritis (71%) compared to those who presented with gastroenteritis but were discharged (41%, p=0.017).

**Table 4.** Clinical characteristics of patients who were discharged compared to those who died

|  | Discharged (N=158) | Died (N=17) | P value |
| --- | --- | --- | --- |
| **Clinical characteristics** |  |  |  |
| Fever | 137 (87%) | 11 (65%) | **0.017** |
| Difficulty breathing | 45 (28%) | 6 (35%) | 0.557 |
| Pneumonia | 38 (24%) | 4 (24%) | 0.962 |
| Gastroenteritis | 64 (41%) | 12 (71%) | **0.017** |
| Sepsis | 68 (43%) | 6 (35%) | 0.539 |
| **Antibiotic in past 7 days** | 61 (39%) | 8 (47%) | 0.498 |
| **Hospital stay** |  |  | 0.616 |
| Mean days (SD) | 9.4 (6.6) | 8.6 (6.4) |  |
| Range | 1.0 - 25.0 | 1.0 - 22.0 |  |

## Discussion

In our study, bacteraemia was highest among children with SAM at 11% despite having the lowest rates of fever and admission diagnosis of sepsis, which are the classical clinical indications for blood culture collection (Supplementary table 1) (8). Mortality (19%) was highest among children with SAM compared to other nutrition groups. Our study highlights the need for blood culture collection on hospital admission for all SAM children at an urban tertiary hospital regardless of their presenting clinical characteristics, as they are at the highest risk of mortality.

The mortality rate among children with SAM found in our study is higher than that in a referral hospital in central Malawi, Kamuzu Central Hospital, where mortality was 10.1% (21). One potential reason for the difference could be because of different geographical locations of the study sites which has an effect on the HIV prevalence with the highest rates in the Southern region (21). HIV is independently associated with death especially in SAM children. In addition, 8.2% (n=54) were lost to follow-up during admission in the other study. The mortality was also higher compared to a study in Ethiopia which assessed bacteraemia in children one to fourteen years of age, and had a mortality of 7.1%(15). This is likely due to the difference in the age of the participants, with our study having a younger population which has a higher risk of mortality. It is important that interventions should target these vulnerable groups.

The overall mortality rate was 10% and over two-thirds of children who died presented with gastroenteritis which encompasses vomiting and diarrhoea. Gastroenteritis was also prevalent in the malnourished population which also had the highest mortality. Diarrhoea is a a risk factor of mortality which worsens with malnutrition and HIV co-infection (28). HIV infection is a risk factor to malnutrition and increases susceptibility to persistent diarrhoea and opportunistic infection, greatly impacting on mortality of hospitalised malnourished children (27). In our study, HIV co-infection was higher in children with SAM and MAM (28% and 29%) compared to NAM (3%) groups.

Malnutrition was present in over half (61%) of the study population and 43% had severe malnutrition. This was higher compared to other countries within the region (6,15,23) and is reflective of the high malnutrition burden in Malawi which has 37% stunting in children under five years of age (24). It is also worth noting that participants that met criteria for NAM still had a mean weight-for-height z score of -0.6 despite having a normal mean MUAC. Less than two-thirds (63%) of children with SAM from the main study had a blood culture collected during admission despite the hospital’s recommendations to have blood culture in all SAM children on admission. Failure to recognise SAM on admission leads to failure to adhere to protocols for management of children with SAM (22). The difficulty of venepuncture in this population due to oedema or severe wasting and parents’ refusal to consent for blood collection also contributed to the lower than targeted blood culture collection rates.

The prevalence of bacteraemia was 11% among children with SAM in our study, higher than those in other nutrition groups. This is comparable to most studies in Sub-Saharan Africa (23,25–27). Mortality in our study was higher in children with than without bacteraemia (38% vs 7%) which aligns with findings in other African studies (5,15,18,25). Fever was less common in children compared to those without bacteraemia (62% vs 86%, p=0.017), possibly because of the depressed immune response among SAM children who accounted for the highest proportion of bacteraemia.

Gram negative bacteria were the predominant isolates from the community acquired infections in children with SAM, including *E. coli, Salmonella Typhimurium* and *E. cloacae*. These enteric pathogens are commonly described in the SAM population, where gut integrity is compromised (27–29). These isolates were resistant to Gentamicin, a first-line antibiotic in our setting. Our results, similar to a recent study in Tanzania (11), but different from older African studies (12,25,30,31), indicate the emerging issue of antibiotic resistance in Sub-Saharan Africa. All isolated pathogens with the exception of *Salmonella Typhimurium* were resistant to Ceftriaxone. Among hospitalised under 5 children at QECH, we documented a rise in resistance to first-line antibiotics from 3.4% to 30.2% between 1998 to 2017 (32). Possible reasons for rising resistance to antibiotics are injudicious use of antibiotics during pre-referral management of patients (over 30% of the patients received antibiotics within 7 days before admission to our study hospital) inappropriate prescribing and excessive use of broad-spectrum antibiotics.

We identified two hospital-acquired bloodstream infections, *K. pneumoniae* and *Candida* species which is similar to a South African study which found *K. pneumoniae, Candida* and *Acinetobacter* as the predominant hospital acquired infections (33). The resistance of *K. pneumoniae* to both first- and second-line antibiotics has been noted in other high and low income settings (11,34–36) and presents a huge challenge with developing effective empirical regimens in settings with limited diagnostics, constrained supply and variety, and rising rates of antimicrobial resistance. Nosocomial infections are also of increasing concern as other studies have shown high mortality and association with prolonged hospital stay (37). Given high rates of resistance shown in the current and previous studies, blood culture should be collected in all SAM patients on admission to ensure administration of appropriate antibiotics, and which can reduce mortality and morbidity in this population.

The main limitations of the study include the small number of children with a blood culture taken/convenience sampling, which limits our ability to generalise. However, this is offset by the strengths of the study, which include the detailed clinical data collection and robust diagnostic microbiology.

Findings will inform the development of evidence-based guidelines. There is need for improved diagnostic capacity in low resource settings to meet the challenge of growing AMR worldwide, combined with translational research, clinical trials and antimicrobial stewardships initiatives modified for implementation in low resource settings, to build an evidence-base to inform management of malnourished children.

## Conclusion

Mortality and bacteraemia rates are highest in hospitalised children with SAM. Despite this, these children do not present with typical clinical features including fever, vomiting and convulsions. Given the high rate of antimicrobial resistance in this setting, a high index of clinical suspicion for infection before starting antibiotics, awareness of local susceptibility patterns and evidence-based antibiotic guidelines are needed to optimise clinical care and antimicrobial stewardship.

## Data Availability

This was a secondary analysis of data from the Childhood Acute Illness Network and Nutrition(CHAIN) network database.

## Acknowledgements

We wish to express our appreciation to all study participants, CHAIN members of staff particularly Isabel Potani, Allison Daniel and Emmanuel Chimwezi; MLW laboratory staff and James Chirombo.

## Supplemental material

**Supplemmentary table 1**. Clinical characteristics and outcomes of SAM patients who had a positive and negative blood culture

