## Supplemental Table 1 for "Childhood malnutrition and bacteraemia at a tertiary hospital in Malawi"

Characteristics and outcomes of SAM patients who had a positive and negative blood culture

|  | No Growth (N=67) | Significant growth (N=8) | p value |
| --- | --- | --- | --- |
| Clinical characteristics |  |  |  |
| Fever | 49 (73%) | 3 (38%) | 0.039 |
| Diarrhoea <14 days | 36 (54%) | 4 (50%) | 0.842 |
| Poor feeding/weight loss | 23 (34%) | 3 (38%) | 0.859 |
| Pneumonia | 16 (24%) | 3 (38%) | 0.403 |
| Sepsis | 21 (31%) | 1 (12%) | 0.269 |
| Antibiotics in the past 7 days | 28 (42%) | 5 (62%) | 0.265 |
| Antibiotics given on admission |  |  |  |
| Benzyl penicillin | 43 (64%) | 4 (50%) | 0.433 |
| Gentamicin | 55 (82%) | 7 (88%) | 0.702 |
| Ceftriaxone | 5 (7%) | 0 (0%) | 0.424 |
| Duration of hospital stay |  |  | 0.794 |
| Mean (SD) | 10.1 (6.9) | 10.9 (7.6) |  |
| Range | 1.0 - 36.0 | 3.0 - 26.0 |  |
| Died | 10 (15%) | 4 (50%) | 0.016 |
